# Dental practice in Pre- COVID19 and Future Perspectives

**DOI:** 10.1101/2020.11.24.20238303

**Authors:** Ashwin Parakkaje Subramanya, MLV Prabhuji

## Abstract

**Objectives:** To assess the trends in dental practice in pre-COVID19 times and future perspectives.

**Methods:** The cross-sectional study was conducted among the dental professionals across India. The snowball and convenience sampling methods were used to ensure maximum participation of the subjects. An online structured questionnaire was created using free-access Google Forms application and was sent to the dental professionals via E-mail, WhatsApp mobile application and through other social media platforms. Initially, a pilot study was conducted among 30 dentists to check for the validity. The questionnaire was administered between the months of May 2020 – July 2020 and was sent to 650 dentists from various parts of the country. Data were compiled and subjected to statistical analysis.

**Results:** The questionnaire was sent to 650 dental professionals. Out of 650, 120 (18.4%) participants took part in the survey. Majority of the respondents had 1 to 3 years of experience in dental practice (n=51, 42.5%). It was noted that majority of the participants were into consultation practice (n=69, 57.5%). 65.8% participants reported that they can withstand upto 6 months from economic view point. Difficulty in paying salaries, reduced incomes is some of the main problems encountered by dental professionals. Increasing the price of treatment, reducing co-workers are a few strategies planned by dental professionals to mitigate the economic burden caused by COVID19 pandemic.

**Conclusion:** Majority of dental professionals in India are dependent on private practice and consultation practice. Due to COVID19, source of income has disrupted for majority of the younger dental professionals.

## Introduction

The Coronavirus disease caused by severe acute respiratory syndrome-coronavirus-2 (SARS-Cov-2 virus) has spread all over the globe. WHO has declared the event as public health emergency of international concern.^1^ The coronavirus infectious disease 2019 (COVID-19) pandemic has led to enormous pressure and burden on health care fraternity around the world. Dentists/dental professionals are considered to be in the very high-risk group for COVID-19 by Occupational Safety and Health Administration (OSHA).^2^ The current pandemic situation has mandated a revision of the existing disease and infection prevention protocols in health care facilities, especially in dental clinics/ hospitals.^3^ However, before the COVID-19 pandemic, dental professionals used masks, gloves, and protective eyewear in routine dental practice, particularly in procedures with dental aerosol production. In this regard, dental professionals were using protective measures even before the emergence of current pandemic situation. Also, there are no reports of respiratory-transmitted diseases, including severe acute respiratory syndrome coronavirus and SARS-CoV-2, in dental professionals or patients in a dental practice.^4^ As per reports it is noted that posterior oropharyngeal saliva samples with COVID19 patients presented with high viral loads shortly after the onset of symptom, which suggests that transmission of virus could take place even when the infected individual is asymptomatic.^5^ It is an evolving situation and there are discrepancies in the evidences available. Hence, it is wise to exercise caution and follow stringent measures to prevent risk of transmission of the disease.

Government of India called for nationwide lockdown to prevent the rapid spread of virus. The method of disease control is based on containment measures and movement control order.^6,7^ Due to the nature of risk of transmission in dental care settings, Dental Council of India directed to provide only emergency dental treatments.^8^ With the nationwide shutdown, dental professionals chose to shut down their practices. To reopen the dental practice, dental practitioners had to redesign and renovate the dental clinic set up in order to prevent risk of transmission and cross contamination. This required purchase of inventory, additional personal protective equipment’s and so on. With all these developments dental practice before COVID-19 pandemic and at present has seen several changes. The aim of the present survey was to assess the nature of dental practice before COVID-19 and future perspectives of dental practitioners across the country.

## Materials and Methods

### Study design

The cross-sectional study was conducted among the dental professionals across India. The snowball and convenience sampling methods were used to ensure maximum participation of the subjects. An online structured questionnaire was created using free-access Google Forms application and was sent to the dental professionals via E-mail, WhatsApp mobile application and through other social media platforms. Initially, a pilot study was conducted among 30 dentists to check for the validity. The questionnaire was administered between the months of May 2020 – July 2020 and was sent to 650 dentists from various parts of the country. The questionnaire consisted of two sections; first section was on basic demographic details, work experience, specialty, work setting and type of dental procedure most often done in their practice; second section consisted of questions on future perspective of dental practice. The questions had multiple choice type answers and questions with multiple answers as well. The study included both Bachelor of Dental Surgery (BDS) and Master of Dental Surgery (MDS) graduates and those who responded to within a 2 week time frame. Figure 1 shows the timeline of the study.

### Ethical considerations

The questionnaire was designed to be anonymous, and informed consent was obtained from every respondent. The data were kept confidential and the results did not identify the respondents personally.

### Statistical Analysis

Statistical analyses were performed using the software Statistical Package for Social Sciences (SPSS) version 22.0 (SPSS, Inc., Chicago, IL, USA). Descriptive statistics were calculated. Pearson’s correlation coefficient was used to investigate the correlation between age, years of experience and some influencing factors like fear of carrying infection to the family, closure of dental clinic until decline of COVID19 cases, endurance to situation from economic point of view and change in practice situations.

## Results

### Demographic details

The questionnaire was sent to 650 dental professionals. Out of 650, 120 (18.4%) participants took part in the survey. The age of participants ranged from 24 years to 66 years. Out of 120 participants, 51(42.5%) participants were male and 69(57.5%) were females. Among the total participants 85 (71.1%) were from Southern states of India, while the rest 35(28.8%) were from other parts of the country.

### Dental practice related data

Majority of the respondents had 1 to 3 years of experience in dental practice (n=51, 42.5%). It was noted that majority of the participants were into consultation practice (n=69, 57.5%) followed by private practitioners (n=51, 42.5%). Most of the participants were general dental practitioners (n=51, 42.5%). Root canal treatment and restorative treatment are the commonly performed treatment followed by dental extraction.

### Future perspectives of Dental Practice

53.3% (n=64) of the participants believed that dental practice will return to normal post-COVID19 pandemic, while the rest 46.7% (n=56) believed it will not. Among those who said that dental practice will not return to normal like pre-COVID19 times (n=26, 46.3%), majority of them were of the opinion that ‘Virus will continue to exist, we will have to live with the virus’ (n=38, 68.9%). Other reasons were ‘Use of Personal Protective Equipment (PPE) shall change the practice’ (n=30, 53.3%) and there will be development of new equipment and instrument to reduce aerosols’ (n=22, 40%).

**According to you what are the disinfection protocols you think should be adapted in dental practice ?**

Air purification - HEPA filter (n=73) 61.3%, UVC disinfection (n=65) 53.8%, Fumigation/ Fogging(n=90) 75%, Surface disinfection using mops and wiping(n=76) 80%, Ventilation ensuring laminar airflow (n=78) 65%, High Volume Suction (n=84) 70%.

**If you have opened your dental practice post-lock down, which among the following treatments are being performed in your clinic ?**

Non-aerosol generating procedures (n=66) 55.3%, Both aerosol and non-aerosol generating procedures (n=33) 27.6%, Only out-patient check up (n=53) 44.7%.

**How long do you think, you can endure this situation from economic point of view?**

1-6 months (n=79) 65.8%, 6-12 months (n=28)23.3%, More than a year(n=13) 10.8%.

**How do you think this situation will change practice from economic point of view?** Difficulty in paying expenses of dental clinic (n=84) 70%, Difficulty in paying co –workers (n=53) 63%, Reduced income (n=105) 87.5%.

**What measures would you consider to run your dental practice from economic point of view ?**

Reduce no. of co-workers (n=43) 36.3%, Reduce the wages (n=12) 10%, Reduce working hours (n=45) 37.5%, Increase the price of treatments (n=86) 71.3%, Depending on alternative source of income (n=54) 45 %.

### Correlation analysis

Relationship between gender, years of dental practice with fear of carrying infection to the family, economic endurance were analyzed using Pearson’s Chi Squared test. There was no significant correlation between gender and fear of carrying infection from dental clinic to the family and also years of dental practice vs fear of carrying infection to home from dental clinic. It was noted that there was statistically significant correlation between years of practicing dentistry vs economic endurance, showing that participants with more years of practicing dentistry had better economic endurance.

## Discussion

The COVID19 pandemic has several implications on dental practice. The present study assessed for nature of dental practice before the COVID19 pandemic and future perspectives. Majority of the surveys have concentrated on knowledge, awareness and practice type of questionnaires on COVID19. The present survey was distinct in terms of assessing the nature of dental practice by dentists before COVID19 and their perspective of future dental practice.

In the survey conducted by DCI it was noted that the prevalence of dental caries was 51.9% in 5-year-old children to as high as 85.0% in adults aged 65–74 years. Dental caries was also reported as the primary cause of edentulism in almost 30% of the senior citizens.^9^ Considering these facts it is relatable that majority of the treatments performed in Indian dental practice is Root canal treatment followed by restorative treatments and dental extraction as noted in present survey. It is reported that around 5 % of the of dentists are employed in public health-care systems.^10^ It was reflected in present survey as well that 6.7% (n=8) dentists reported to be working at government setup. Majority of the dental professionals in India are dependent on private practice (42.5%) and consultation practice (57.5%).

In a survey on perceived COVID-19 risk in dentistry and the possible Use of rapid tests, majority of the dentists were of the opinion that they prefer patients to have in-office rapid test for COVID-19 before treatment (79.29%, n=555/700).^11^ Our results on the question ‘Do you think patients should undergo Covid-19 testing before dental treatment ?’, majority of the participants were of the opinion that patients should undergo COVID19 testing before dental treatment (Yes - 58.3%, n=70; Maybe −33.3%, n=40). On the question on ‘Do you have fear that you could carry infection from your practice to your family ?’, majority of the participants reported yes’(96.3%,n=115). Similar results were reported in a previous survey. Participants perceived a very high risk of COVID-19 contamination during the dental procedure.^11^ Thus the opinion of majority of dental professionals, ‘patients should undergo COVID19 testing before the treatment’ can be related to the fear of carrying infection to home and also regarding risk of cross-contamination in dental practice. Although universal precautions and emphasis on personal protection equipment are heightened due to COVID19, the possible risk of spread cannot be fully ruled out. However, there are mixed opinion on this issue. In their report by Niraj et al, it is reported that small majority (57.8%) perceived that they like to request COVID-19 test results from all patients prior to any aerosol-generating treatment procedures, while the rest (40.0%) of the respondents wished to request the test only from symptomatic patients.^12^ Regarding the question on closure of dental practice until pandemic ends, only 40% of the respondents said ‘yes’. In a survey by Niraj et al, one half of the respondents (54.3%) were not confident, and about a third (35.7%) hesitant to commence their post-pandemic dental practices. And the reasons cited were risk of carrying infection and additional financial burden to additional infection control measures.^12^

It is well known that COVID19 has led to economical hardships to dental professionals as well. In the present survey it was noted that majority of the participants said they can endure the economic impact only up to 6 months. It was also noted that this opinion was majorly among the younger dental professionals with less years of practice. Also dental professionals expressed their concern on difficulty in paying the expenses, difficulty in paying co-workers and paying wages, and also reduced income. This was also reported by Ahmadi et al that majority of (97%) of dental professionals reported that they encountered a decrease in their financial income since the pandemic.^13^ In a survey conducted at UK, it was reported that volume of inpatient per day was reduced by 74% due to the pandemic situation.^14^ To overcome the economic impact, majority of dental professionals are considering following measures: Increase the price of treatment and depend on alternative source of income. Other measures being thought by dentists are reducing the no. of coworkers and also to reduce the working hours. Governments in few countries have considered special economic packages to dental professionals to overcome the economic burden.^15^ It is desirable that Government of India also considers a package for dental professionals.

## Conclusion

Majority of dental professionals in India are dependent on private practice and consultation practice. Due to COVID19, source of income has disrupted for majority of the younger dental professionals. There is mixed opinion regarding COVID19 test prior to dental treatments. Increasing the price of treatment, depending on alternative source of income are certain measures are what dentists are looking forward.

## Data Availability

All the data related to this manuscript is available with authors.

---

**Table 1:**
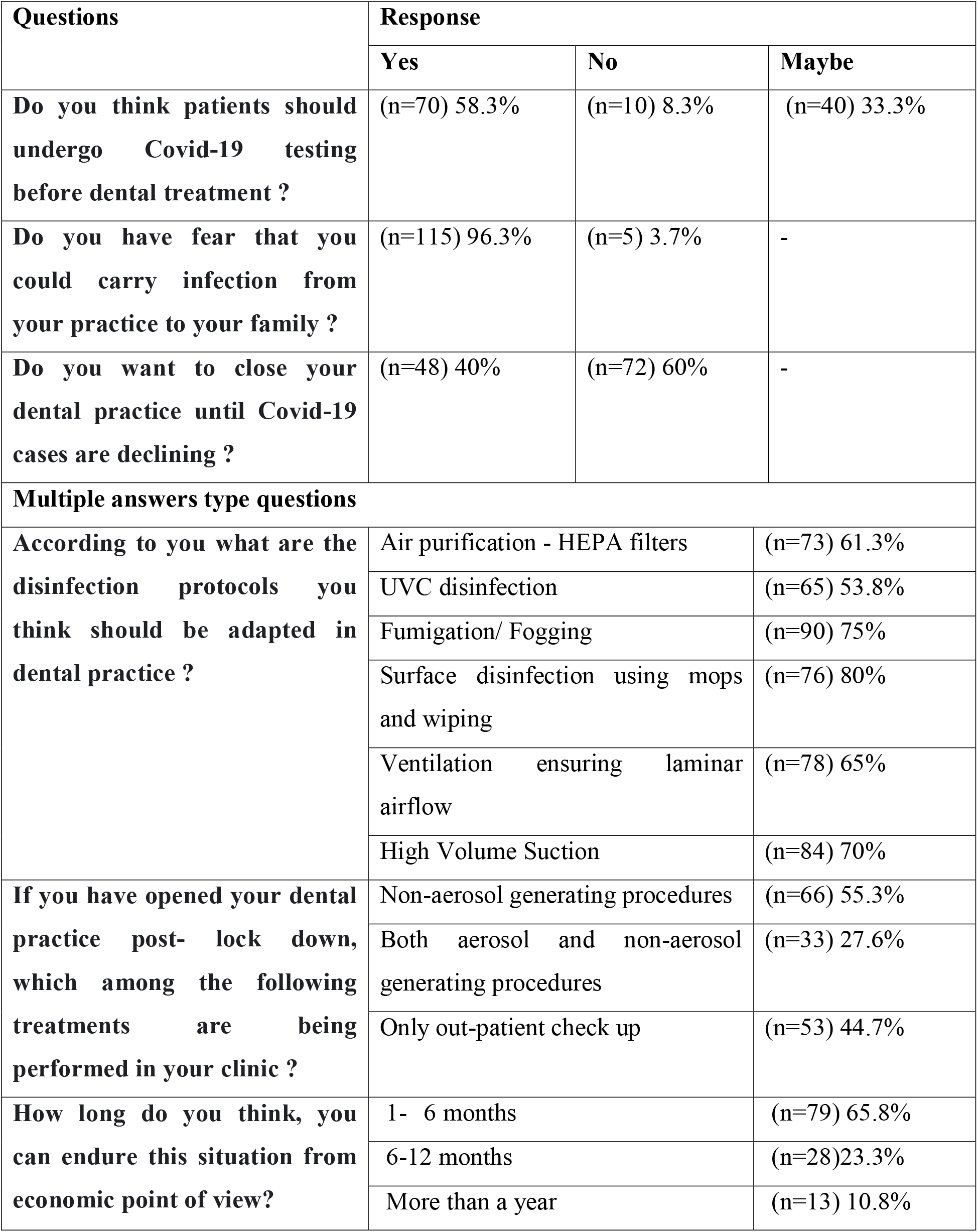
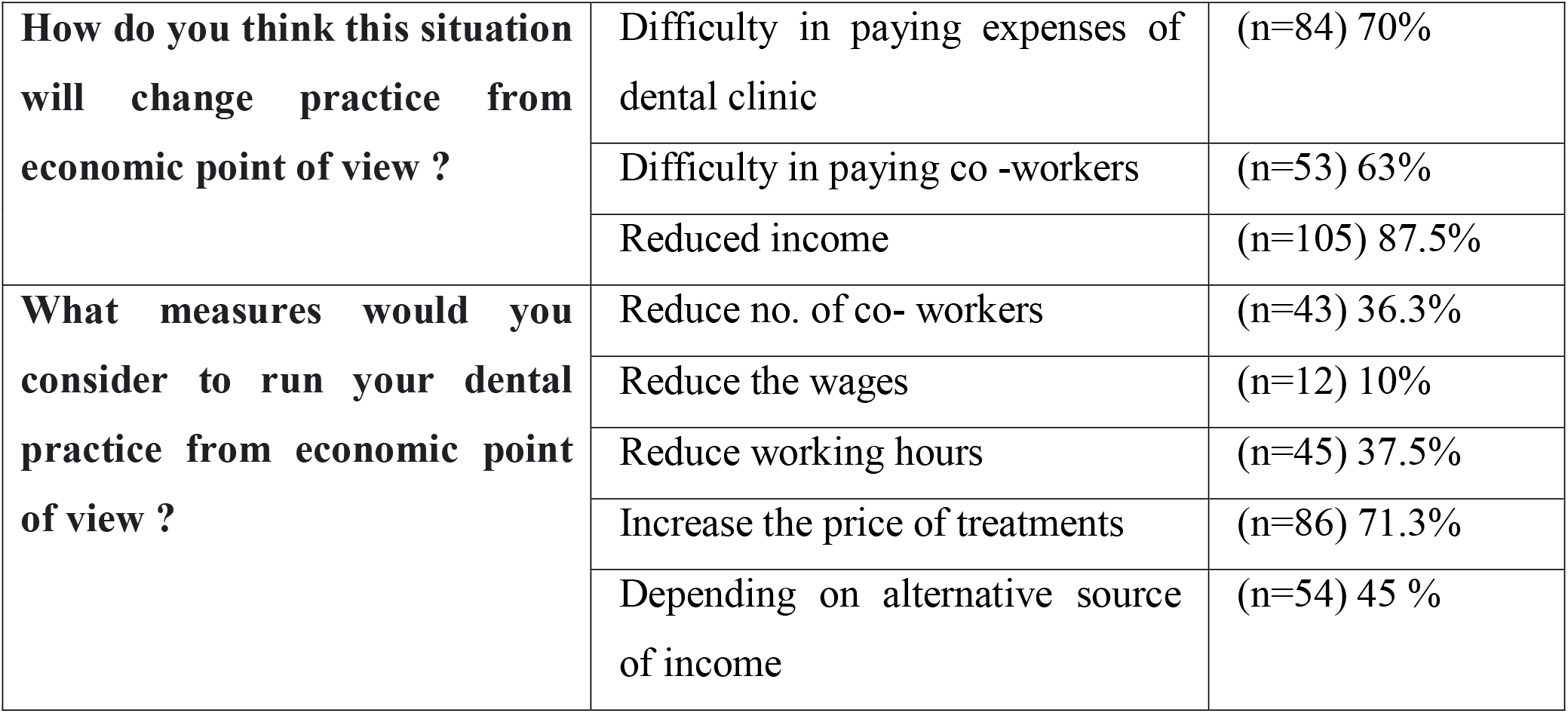
Reponses for questions on future perspective of dental practice due to COVID19 pandemic.

